# Screening for SARS-CoV-2 infections in daycare facilities for children in a large city in Germany

**DOI:** 10.1101/2021.02.26.21252510

**Authors:** Nadine Lübke, Anna-Kathrin Schupp, Renate Bredahl, Ursula Kraus, Sandra Hauka, Marcel Andrée, Lutz Ehlkes, Thomas Klein, Alexandra Graupner, Johannes Horn, Ralph Brinks, Klaus Göbels, Ortwin Adams, Jörg Timm

**Author notes:** **Corresponding author:** Joerg Timm, Institute of Virology, Heinrich-Heine-Universität, Universitätsklinikum Düsseldorf, Universitätsstraße 1, 40225 Düsseldorf. **Alternative corresponding author:** Dr. Nadine Lübke, Institute of Virology, Heinrich-Heine-Universität, Universitätsklinikum Düsseldorf, Universitätsstraße 1, 40225 Düsseldorf.

## Abstract

**Background:** The role of pre-school children as a source and distributor of SARS-CoV-2 infections is still unclear. Daycare facilities that care particularly for young children with limited hygiene measures may contribute to the infection dynamics during the pandemic. The aim of this study was to implement and evaluate a voluntary SARS-CoV-2 screening program in daycare facilities.

**Methods:** The study was conducted over a period of 4 weeks, from June 10^th^ to July 7^th^ 2020. The aim was to screen a representative group of 5000 individuals (children and staff at a ratio 3:1) attending daycare facilities in Düsseldorf, North Rhine-Westphalia. Tests were performed twice per week with oral rinsing water as sample material for the detection of SARS-CoV-2-RNA by molecular pool testing.

**Results:** A total number of 5210 participants (75.9% children and 24.1% staff) from 115 day care centers participated in the study. Of a total of 34,068 returned samples (81.7%) during the study period, only one SARS-CoV-2 infection of a child was detected in the study cohort with one likely secondary infection within the daycare facility. Of note, during the study phase, no increase of SARS-CoV-2 infections was observed in daycare center compared to the overall incidence in Düsseldorf.

**Conclusions:** A voluntary screening program for SARS-CoV-2 infections could successfully be implemented in daycare facilities. Although the low overall incidence during the study period precludes firm conclusions, there was no evidence for increased transmission in children attending daycare facilities compared to the general population of Düsseldorf.

**Summary:** SARS-CoV-2 screening programs in daycare facilities may help to detect asymptomatic infections at an early stage and thereby support containment. Here, a large screening study was evaluated suggesting similar infection rates in daycare facilities compared to the general population.

## Introduction

Starting in December 2019, a new coronavirus (SARS-CoV-2) emerging from China has caused a global pandemic [1]. In response, measures were implemented with the aim to limit the number of contacts and thus preventing further viral spread. One measure was the closing of schools and day care centers for children. Although children have a low risk of severe illness upon SARS-CoV-2 infection [3-5[2-4], it was believed that children were a relevant source of new infections, as observed for other infectious diseases of the respiratory tract [5, 6]. Moreover, infections may be frequently asymptomatic in children and thus contact restrictions for symptomatic cases are not sufficient [7]. At the same time, children are considered an important link to risk groups, as cross-generational contacts are common among them [8].

Against this background, reports have received a great attention, stating that children may not only have a milder course of the disease but, in addition, infections occur less frequently. This assumption was based on observations from population-based studies showing a lower Anti-SARS-CoV-2 seroprevalence in children [9]. Children were also less likely to be infected in case studies of households [8]. However, biased case finding among children in these studies may have contributed to the observation. In turn, when the concentration of viral RNA in test material of infected individuals from different age groups was compared [10], no differences were found between children at preschool age compared to the other age groups. Moreover, in the REACT study in the UK, the frequency of proven infections in children at the age group between 5 and 12 years was in the same range compared to older age groups [11].

Although opening daycare facilities for children in preschool age is of social interest due to the negative impacts of closed daycare centers on child development [12], it may be associated with an increased transmission risk. Adherence to general hygiene rules, e.g. distancing or wearing masks, is difficult to comply with in this age group. The data basis for assessing the importance of children as a source of infection is insufficient for clear recommendations. Therefore, the aim of the study was to implement and evaluate a voluntary screening strategy for monitoring of SARS-CoV-2 infections concomitant to the re-opening of the day care centers.

## Methods

### Recruitment of study participants

On June 8^th^, 2020, the regular operation of daycare facilities for children was resumed in the state of North Rhine-Westphalia, Germany. All non-symptomatic children without direct contacts to infected people were again entitled to a place day care. The starting date for the voluntary screening program was set to June 10^th^ for collection of the first sample. To reach out to the families and childcare workers the Youth Welfare Service of Düsseldorf contacted daycare facilities with information on the study. At the same time, hotlines and e-mail accounts were set up, to which questions about the study could be addressed. All interested persons were reported to the Youth Welfare Service, where a study group of 5000 individuals from spatially and socially representative daycare centers was selected for the voluntary screening program. The selection of participating daycare centers in the study was based on the ratio of voluntary participants of children to staff (ratio 3:1) and according to a representative distribution of the facilities depending on the different social areas in Düsseldorf. All participants signed a consent form and the study was approved by the ethics committee of the Medical Faculty of the University of Düsseldorf. The allocation to social areas follows the social area classification of the city of Düsseldorf and is available from the Office for Statistics and Elections [13]. According to this version, five levels of social burden are distinguished based on the need for social action in a residential area (“very low” = 1, “low” = 2, “medium” = 3, “high” = 4 and “very high” = 5).

### Study procedure

During the study period of 4 weeks, the participants were meant to be tested twice a week for a SARS-CoV-2 infection. Therefore, eight sample vials with individualized barcodes were prepared at the Institute of Virology and provided to the study participants, together with instructions for the sample collection procedure. All participating day care facilities were divided into two groups, which were scheduled for sample collection either on Monday and Thursday or on Tuesday and Friday. For the detection of SARS-CoV-2, “spit samples” were used. In detail, 10 ml of water should be rinsed in the mouth in the morning before brushing the teeth and before the first food and drink intake and then spat into the sample container. The samples were collected at the day care center and transported collectively to the Institute of Virology. Here, pools of 20 samples were prepared and tested using a cobas^®^ SARS-CoV-2 assay on a Roche cobas 6800 system. For pools that tested positive all individual samples in the pool were tested again.

### Monitoring of SARS-CoV-2 infections in Düsseldorf during the study period

The total number of SARS-CoV-2 infections during the study period was deducted from the reporting files of the Public Health Authority of Düsseldorf, which is based on the legal obligation to report positive SARS-CoV-2-RNA test results. Accordingly, the number of infections per 1,000 inhabitants or, in reference to day care facilities for children, the number of infections per 1,000 children was calculated. The number of inhabitants in Düsseldorf was assumed to be 612,000. The total number of children in care during the study period was derived from the daily statistics of the Youth Wellfare Service. The number of children in study facilities not participating in the study was calculated as the number of total children in care during the study period minus the number of children participating in the study. Comparisons between the numbers of infections in different groups were calculated by Fisher’s exact test and confidence intervals (95%) were calculated according to the method by Clopper-Pearson.

## Results

### Study population

Out of a total of contacted 364 daycare centers, 314 facilities with a total of 10,289 individuals (7471 children and 2818 childcare workers) responded and declared their willingness to participate in the study. Based on criteria as outlined in figure 1, 115 facilities with a total of 5232 study participants were selected as a representative study group. Consent was withdrawn by 22 persons, resulting in a total of 5210 participants (3955 children and 1255 staff) in the study. In the 115 facilities selected for the study a total 6606 attended the daycare facility during the study period. The study population thus comprises 59.9% of the children in care at the facilities during the study period (ranging from 32.8% to 100% per facility) or in turn, 40.1% of the attending children in participating day care facilities were not participating in the study. The age distribution and the social affiliation according to the level of social burden of the participating children in the study are shown in Figure 2. Not all daycare facilities accept children at the age <3 years, accordingly, the proportion of children under the age of three is low (5.6%). The social affiliation of children participating in the study reflects the relative frequency of children in the defined social areas in the city of Düsseldorf (Figure 2B).

**Figure 1:**
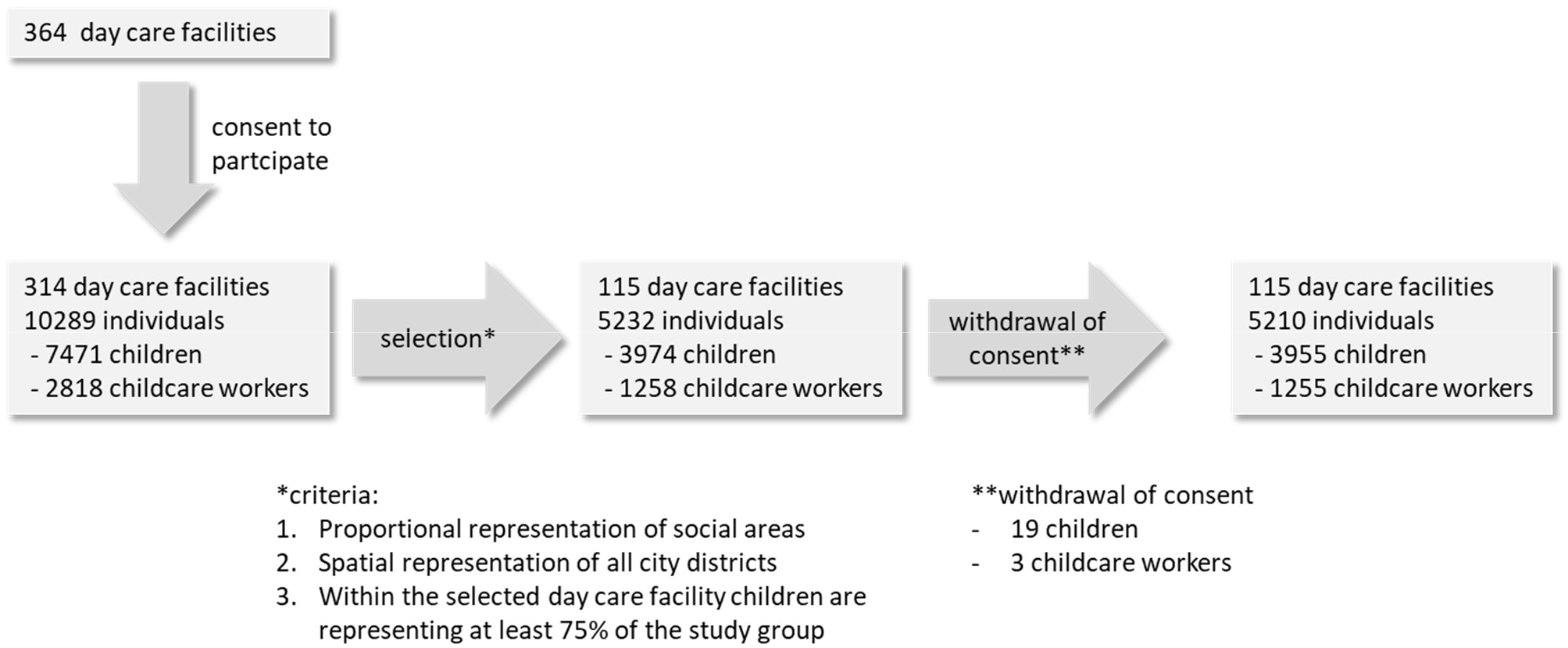
Recruitment of study participants

**Figure 2:**
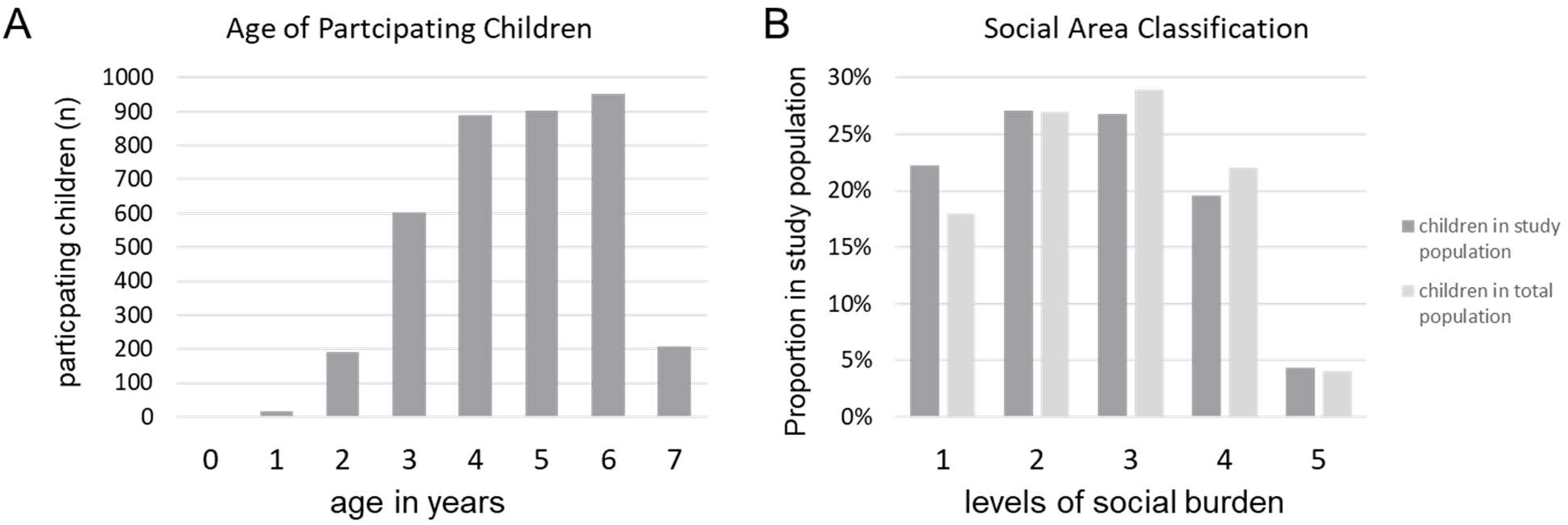
(A) Age distribution of the participating children. (B) Social affiliation of children participating in the study and of children from the total population in the defined social areas in the city of Düsseldorf. The level of social burden is based on the need for social action in defined residential areas (“very low” = 1, “low” = 2, “medium” = 3, “high” = 4 and “very high” = 5).

### Sample return

Samples were collected and tested for SARS-CoV-2 twice a week in two groups from June 10^th^ to July 7^th^. Of the total 5210 individuals who participated in the study, 96.4% (5025 individuals, including 3765 children and 1260 employees) provided at least one sample. Of the 41,680 sample vials distributed, 34,068 (81.7%) were returned to the Institute of Virology for testing. 2076 study participants (39.8%) returned all eight samples (Figure 3). The number of returned samples decreased continuously over time from an initial 89.6% to 71.8%. Of note, in some facilities summer break was starting June 29^th^. Overall, the proportion of returned samples did not differ by the different social areas (ranging from 81.2% to 85.9%; data not shown).The percentage of returned samples among employees was 83.9%.

**Figure 3:**
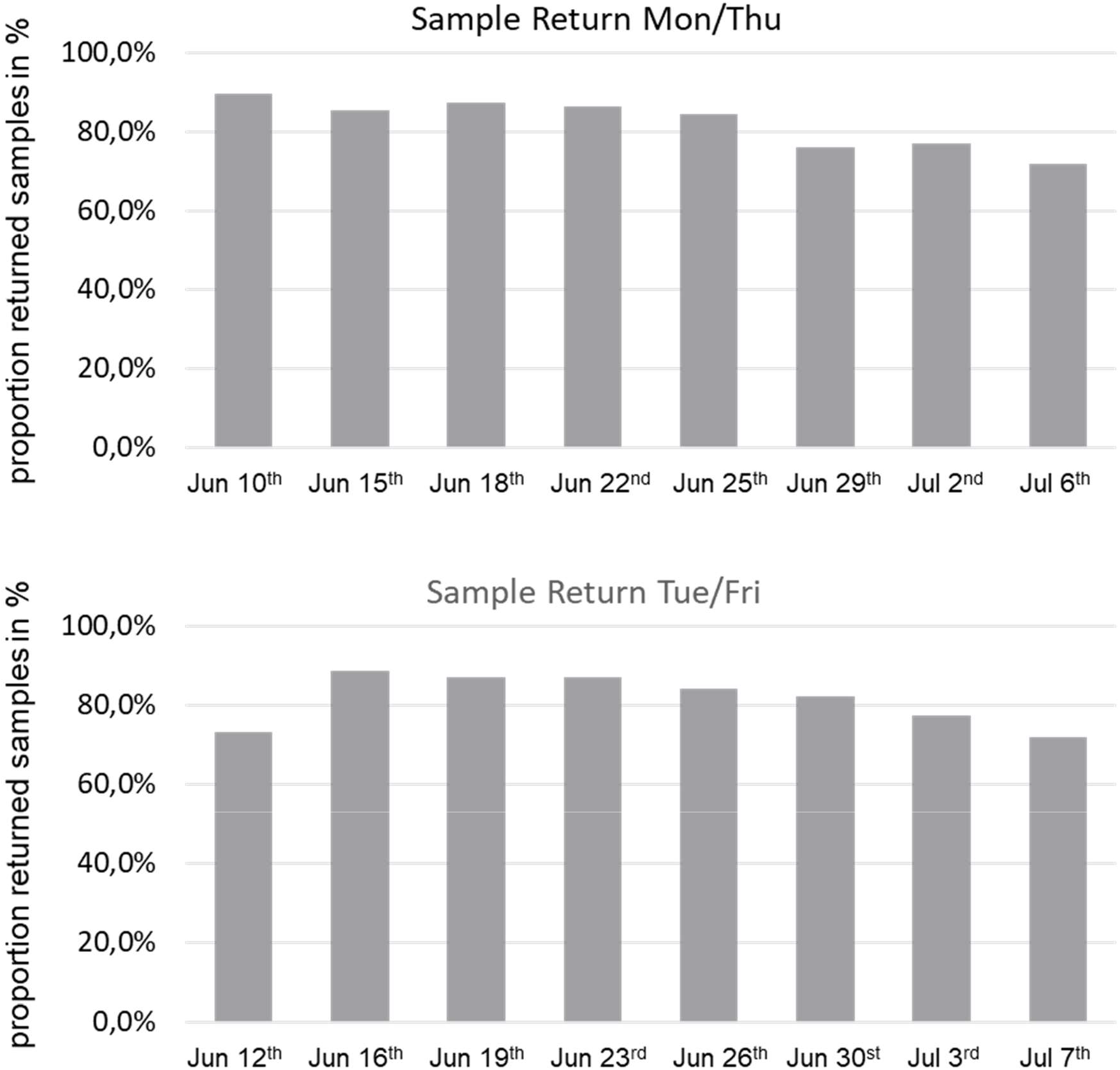
Relative numbers of returned samples over the study period.

### SARS-CoV-2 Infections reported to the public health department of Düsseldorf

During the study period 501 infections were reported to the Public Health Authority of Düsseldorf. Thirty-two reported infections were among preschool-aged children (born after 9/30/2013). Of the 32 infections in preschool-aged children, 16 were registered in a daycare facility for children. During the study period 15,265 of the children registered at day care were actually visiting the facilities in Düsseldorf. Accordingly, the proportion of SARS-CoV-2 infections in children in day care facilities in Düsseldorf was 1.05 per 1,000 children (95% CI 0.60-1.70; Figure 4) during the study period. The frequency of reported SARS-CoV-2 infections in day care during the study period was not significantly different from the prevalence of reported infections in the general population of Düsseldorf (0.81 infections per 1000 inhabitants, 95% CI 0.74-0.89, p=0.313 by Fisher’s exact test; Figure 4).

**Figure 4:**
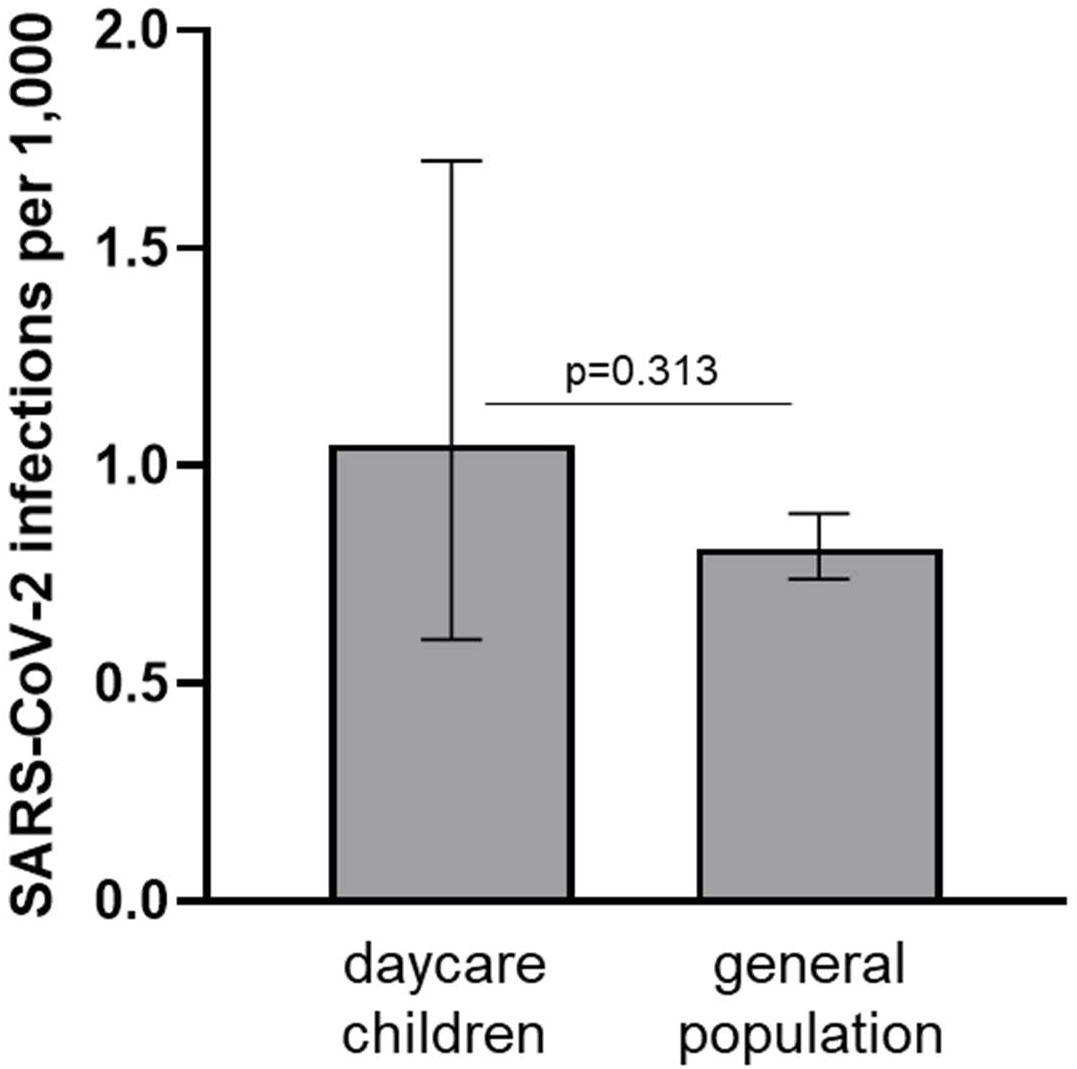
Number of Sars-CoV-2 infections over the study period in the population of the city of Düsseldorf and in children attending a day care center. The frequency of infections per 1,000 children or 1,000 inhabitants with 95% CI according to Clopper-Pearson is shown. The p-value was calculated by Fisher’s exact test.

### SARS-CoV-2 Infections in day care facilities participating in the study

Based on the reports to the Public Health Authority, a total of 10 infections (eight children and two childcare workers) could be assigned to facilities participating in the study (Table 1). Notably, only one infection was identified in a child as part of the study group, whereas seven infections were identified in children who did not participate in the study. Accordingly, in day care facilities participating in the study the incidence was significantly lower in the study group (0.25 infections per 1,000 children; 95% CI 0.01-1.41) compared to children who did not participate in the study (2.64 infections per 1,000 children, 95% CI 1.06-5.43, p=0.0087 by Fisher’s exact test; figure 5). Reasons for SARS-CoV-2 testing of children not participating in the study were recent contact to confirmed infections and/or symptoms consistent with SARS-CoV-2 infections (Table 1).

**Table 1:** SARS-CoV-2 infections in daycare centers in the study period from June 10^th^ to July 7^th^ 2020.

| case # | age | day care center | social area | reason for testing | contacts in day care | infections in day care contacts | additional infections in household | comment |
| --- | --- | --- | --- | --- | --- | --- | --- | --- |
| 1 | 6 years | RO | 4 | study | 20 | 1 | 2 | index case most likely mother |
| 2 | 5 years | RO | 4 | contact | none | - | 3 | contact to case #1 five days prior to testing |
| 3 | 6 years | AL | 4 | contact | 22 | none | 3 | index case most likely father |
| 4 | 6 years | SB | 3 | contact | 19 | none | none | same contact as case #5 outside the day care center |
| 5 | 5 years | SB | 3 | contact | none | - | none | same contact as case #4 outside the day care center |
| 6 | 5 years | BL | 1 | symptoms | 66 | none | none | quarantine of the entire day care facility |
| 7 | 4 years | BR | 2 | contact | none | - | 2 | index case most likely father |
| 8 | 2 years | ZA | 2 | symptoms | 22 | none | none |  |
| 9 | 43 years | SB | 3 | unknown | none | - | none |  |
| 10 | unknown | PS | 2 | unknown | none | - | unknown |  |

**Figure 5:**
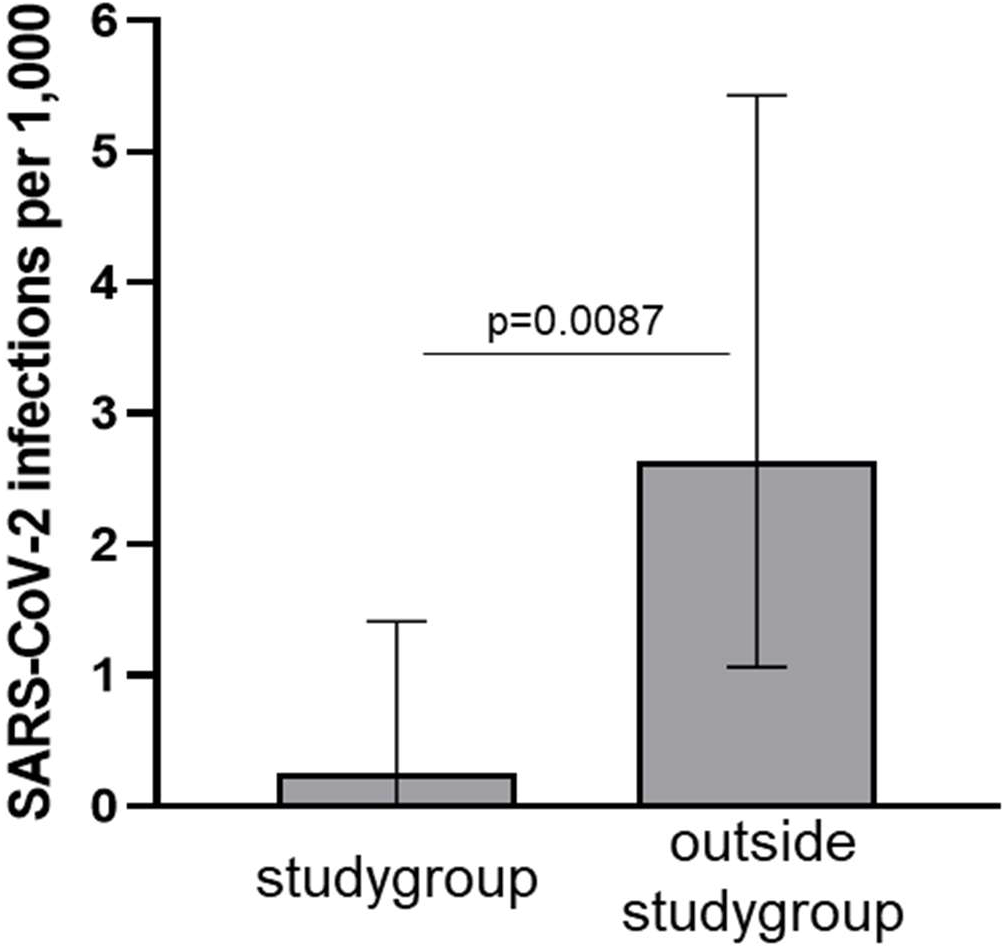
Sars-CoV-2 infections inside and outside the study group in children attending day care centers in the city of Düsseldorf. The frequency of infections per 1,000 children with 95% CI according to Clopper-Pearson is shown. The p-value was calculated by Fisher’s exact test.

The single infection detected within the study group was a 6-year-old child who tested positive on June 15^th^, 2020. The child was asymptomatic at the time of the positive test result but then developed diarrhea the next day. The mother herself had symptoms of a common cold and dysosmia starting four days before the child was tested positive. After becoming aware of the infection in the child, the mother and a sibling were examined and also tested positive for SARS-CoV-2. The sibling had developed mild symptoms with a slightly elevated temperature one day later. All three cases had a mild disease course. For all detected infections the Public Health Authority investigated possible contacts within and outside the daycare facilities. All contacts participating in the study were offered to continue regular testing within the study, and all contacts not participating in the study were offered testing by the public health department immediately and again on day 7. Upon testing of these contacts one additional infection was detected in a 5-year old child who had contact in the childcare group 10 days earlier to the case detected in the study group. According to the timeline of events transmission within the daycare facility seemed most likely.

## Discussion

The surveillance study for SARS-CoV-2 infections in daycare facilities for children in Düsseldorf was conducted with the aim to determine the incidence of new infections after daycare centers reopened with minimized contact between care groups. During the study period from June 10^th^ to July 7^th^, 2020, SARS-CoV-2 incidence was low in Germany, yet there was a high level of interest from daycare facilities and willingness to participate in the study. The high response rate of guardians and childcare workers allowed a selection of facilities to create a representative study population for the city of Düsseldorf. The return of samples was high (averaging over 80%) indicating a high interest and feasibility of infection surveillance in the context of childcare facilities.

The low infection numbers at this time are also reflected in the low number of observed infections in the study, with only one infection detected after testing 34,068 samples from 5,210 participants within the study and nine additional infections (seven children and two childcare workers) detected outside the study population in facilities participating in the study. This low prevalence during the summer in 2020 was also seen in a similar study performed in the state of Hessen (Germany) [14]. Nevertheless, the low case numbers during the study period also suggest that during a low incidence period, transmissions are rare in the context of day care centers, which is in line with other studies [15-18]. Notably, in regions with higher SARS-CoV-2 burden direct transmissions of SARS-CoV-2 infections in day care centers were more common and transmission chains of SARS-CoV-2 could be observed within the families of the infected children [19, 20]. In our study, we identified one likely child-to-child transmission within the day care facility. Unfortunately, the samples were not available for viral sequencing, however, the timeline of events was highly suggestive for transmission during day care, because this was the only contact reported and no other plausible sources for infection could be identified. Importantly, the child receiving the SARS-CoV-2 infection transmitted to one additional household contact suggesting also here a transmission chain with at least five cases from two households.

It is notable that, despite regular testing, significantly fewer infections were identified within the study population compared to children outside the study group. All infections outside the study group were identified by cause-related testing, when children reported symptoms consistent with SARS-CoV-2 infection or - more frequently - were tested after contact to confirmed cases with SARS-CoV-2 infection. The exact reasons for this substantially lower case finding upon regular screening of the study group compared to cause-related testing are not clear, but have also been observed in other studies [14]. One possible explanation may be a lower test sensitivity when unsupervised “spit samples” were used as sample material and tested in pools, although other studies have already shown that pharyngeal rinse water is considered as suitable material for detection of respiratory pathogens [21-23]. The advantage of oral rinse water is the less invasive procedure which is more appropriate for a regular screening. This seems to be an important aspect in the context of frequent sampling of preschool-aged children. In our own observations and also in smaller case series, SARS-CoV-2 RNA was detected in oral rinse water during the early phase of infection [24]. Saliva has also recently been suggested as a material that can be used to reliably test for SARS-CoV-2 [25]. In synopsis of the data, the use of saliva samples or even spit samples for monitoring incident SARS-CoV-2 infections in daycare centers seemed appropriate. By sampling twice per week, it was hypothesized, that individuals with recent infection and high viral shedding will be detected by a sensitive molecular testing system.

An alternative reason for the higher number of infections outside the study population is a possible selection bias in the study group. It seems plausible that a voluntary screening program in daycare facilities preferentially reached families and childcare workers who are well aware of infection risks. It is conceivable that the awareness of the infection risks differed between the families from the study group and families who did not consent. The lack of consent may be due to various reasons, but could be associated with a perception of a rather low risk of infection. On the other hand, the risk of infection may be perceived as higher among study-participating families, associated with more consistent adherence to preventive measures. Although no systematic evaluation of the reasons for participating in the study was performed, a higher sense of security by regular testing was frequently reported as a reason for participation in the study.

Infections were independently reported to the Public Health Authority and evaluated. A total of 501 infections were reported in the four-week period of which 16 children were attending a day care facility. Because the number of children in care is reported on a daily basis, the number of new infections per 1,000 children attending a day care facility during the study period can be estimated with high reliability. The result of 1.05 infections per 1,000 children is in the same range and not significantly different from the observed 0.81 infections per 1,000 inhabitants of the city of Düsseldorf. Thus, the frequency of infections in daycare centers seems to reflect the general infection situation in Düsseldorf at this time. Accordingly, there is no indication from these data that infections in children in day care centers are less frequent, although there is considerable statistical uncertainty given the small number of cases.

In conclusion, a voluntary screening for SARS-CoV-2 infections in daycare facilities for children was successfully implemented allowing detection of SARS-CoV-2 infections. The detection of infections was lower in the study group participating in the voluntary screening compared to cause-related testing outside the study population. Although the reasons are not clear, differences in the test sensitivity or a selection bias in the study population for a voluntary screening program may explain this observation. For the observed study period, we found no evidence for a higher rate of transmission in daycare facilities for children compared to the general population. However, it has to be highlighted that the study was performed during a low incidence period in Germany in the summer of 2020, which is a limiting factor. Nevertheless, this form of screening program can support early detection of asymptomatic cases in daycare centers. Early isolation of cases may therefore enhance infection containment in a setting where established hygiene measures are difficult to implement and helps maintaining regular operations in the daycare facilities.

## Data Availability

There is no supplementary material. All data is available in the manuskript.

## Acknowledgement

The authors would like to thank the members of the research team of the Institute of Virology as well as the student assistants for their support in sample preparation and testing.

